# The beneficial effect of sleep on behavioral health problems in youth is disrupted by prenatal cannabis exposure: A causal random forest analysis of ABCD data

**DOI:** 10.1101/2022.05.12.22275012

**Authors:** Philip A. Spechler, Roman M. Gutierrez, Susan F. Tapert, Wesley K. Thompson, Martin P. Paulus

## Abstract

**Importance:** Declining risk perceptions and touted medicinal claims about cannabis are likely related to the increasing prevalence of cannabis use during pregnancy. Yet, it remains unclear if prenatal cannabis exposure yields lasting neurodevelopmental alterations in children, and which facets of their behavioral health might be affected.

**Objective:** To determine if prenatal cannabis exposure moderates the relationship between changing sleep hours on changing mood and behavioral problems in youth.

**Design:** Causal random forest analysis of ABCD cohort data using baseline (ages 9-10) and year-1 follow up information.

**Setting:** 11,875 children and their parents were assessed at 21 acquisition sites across the United States between July 2016 to October 2018.

**Participants:** Participants with prenatal drug use data were included, and 94% of the baseline sample also had year-1 follow up data.

**Exposures:** A change in sleep hours from baseline to year-1 was conceptualized as the dimensional “treatment” variable, and self-reported drug uses of the biological mother were conceptualized as moderators of that “treatment”.

**Main Outcomes and Measures:** A change in internalizing (anxious/depressed mood) and externalizing (disruptive behavior) problems from baseline to year-1 were the two outcome measures. Sociodemographic and other prenatal drug exposures were included as covariates. Given the observational nature of the multi-site ABCD study, all hypotheses tested here were formulated after data collection.

**Results:** There were n=9,826 children (4,663 female) included in analyses, of which n=605 (303 female) had any prenatal cannabis exposure. On average, internalizing problems largely remained stable from baseline (*M*=48.54, *SD*=10.56) to year-1 (*M*=48.75, *SD*=10.64)(*t*_9,825_=2.40, *p*=.016), as did externalizing problems (baseline *M*=45.64, *SD*=10.21; year-1 *M*=45.23, *SD*=10.09) (*t*_9,825_=5.44, *p*<.001). Significant average treatment effects indicated that participants with more sleep hours reported less internalizing (ATE=-.35, *SE*=.08, *p*=.003) and externalizing (ATE=-.28, *SE*=.07, *p*=.028) problems over time. However, a significant heterogeneous treatment effect by prenatal cannabis exposure (conditional-ATE=.92, *SE*=.36, *p*=.011) was found for the internalizing model, such that participants with this exposure did not show a beneficial effect of changing sleep on changing mood (*B*=.06, *SE*=.25). This finding was specific to cannabis, as no such effect was found for any sociodemographics or prenatal alcohol or tobacco exposures for the internalizing or externalizing models.

**Conclusions and Relevance:** This study uncovers an actionable target (sleep) to improve mood and behavioral problems in typically developing children not prenatally exposed to cannabis. However, changing sleep may not have a similar effect for youth prenatally exposed to cannabis. Given the importance of the endocannabinoid system in regulating perinatal neurodevelopment and sleep, these findings suggest that cannabis exposure may interact with those processes to diminish the beneficial effects of sleep on mood in children, and thus calls into question the safety of cannabis use during pregnancy.

**KEY POINTS:** *Question:* Do children exposed to cannabis during pregnancy exhibit differences in the effect of sleep on their mood and behavior?

*Findings:* Causal inference analyses of baseline (ages 9-10) and year-1 follow up data of the Adolescent Brain Cognitive Development™ (ABCD) study (N=9,826) suggested that increasing sleep hours lowered mood and behavioral problems in children without prenatal cannabis exposures, however, children with exposures did not exhibit similar beneficial effects of sleep on their mood. This finding was specific to cannabis, as other drug use and sociodemographic information did not influence treatment effects.

**Meaning:** Prenatal cannabis exposure likely interferes with neurodevelopmental processes related to sleep, and these differences persist into at least early adolescence to alter the beneficial effects of sleep on mood.

## INTRODUCTION

Healthy sleep is an essential part of child and adolescent development. An extensive literature highlights the importance of sleep on physiological, neurological, and emotional development (see Tarokh and colleagues for review)^1^. At the same time, deficits in sleep have been associated with disadvantages in each of these domains. For instance, poor sleep has been linked to the development of behavioral health issues like internalizing and externalizing problems^2^, with mounting evidence supporting the directionality that sleep disturbance is a precursor to future mood and behavioral disruption in youth^3,4^.

Various exposures and environmental factors can have profound effects on sleep, which, in turn, might also affect mood and behavior. For example, exogenous cannabinoids like tetrahydrocannabinol (THC) readily cross the placenta and interacts with the fetal endogenous cannabinoid (endocannabinoid) system, which is heavily involved in orchestrating elements of neurodevelopment^5^. The endocannabinoid system is also implicated in regulating neural systems for sleep activity^6^, and may be vulnerable to interference by exogenous cannabinoids (e.g., THC) during development. Several studies suggest that, relative to children without prenatal cannabis exposures, those *with* exposures exhibit poor sleep features including more sleep disturbances (i.e., motility and arousal) and neural waveform differences during sleep-cycles as measured via neonatal electroencephalography (EEG)^7,8^. This pattern of poor sleep following prenatal cannabis exposure appears to persist at 3-years^9^, and into late childhood^10^. As prenatal cannabis use may disrupt the child’s prenatal brain development and influence their sleep systems, children with these exposures may be less equipped to achieve a similar level of sleep quality relative to non-exposed children.

A recent report by Paul and colleagues analyzed data from the Adolescent Brain Cognitive Development™ (ABCD) Study^11^ and found that prenatal cannabis exposure was correlated with higher instances of internalizing and externalizing problems in children at baseline (age 9-10)^12^. Considering the literature highlighting the importance of sleep on mood and behavior in healthy children^1^, and other studies finding sleep disadvantages in children with prenatal exposure, it was therefore hypothesized that an effect of sleep on mental health outcomes would vary by prenatal cannabis exposure status (i.e., moderation).

A rigorous examination of the causal effect of modifying sleep on child mental health outcomes would require a randomized controlled trial, which is impractical and prohibitively expensive. Alternatively, one can begin interrogating causal processes by applying statistical causal inference techniques to observational data^13^. Given the large sample and longitudinal nature of the ABCD Study^11^, this dataset is well suited for causal inference analyses. While inferring causality from observational data is challenging due to the possible influence of unobserved confounders, imperfect measurement of observed confounders, and reciprocal relationships/reverse causality^14^, one may begin to make causal inferences by analyzing data under the assumptions of the potential outcomes framework^13^. This framework infers causality by comparing outcomes between similar participants who either did or did not experience a treatment/exposure (e.g., active drug vs. placebo), and can be extended beyond categorical levels to study dimensional treatments or exposures.

In the present study, a change in sleep hours between baseline and year-1 follow up was conceptualized as a dimensional “treatment” measure to infer its effect on the degree of change in internalizing or externalizing problems (outcome measures). As the goal was to examine a *modifiable* behavioral feature (sleep) potentially influencing child health outcomes, prenatal cannabis exposure was not modelled as the treatment measure because its modification is impossible in this sample.

A causal random forest (CRF) model^15^ was implemented to test our hypotheses. This approach uses causal decision trees to find highly similar participants given a set of covariates (analogous to a matching procedure), and then examines differences in outcomes among those participants. Assuming that all variables influencing changes in sleep hours and mental health outcomes have been measured and accounted for (“unconfoundedness assumption”), the CRF approach has been shown to make valid statistical inferences^15^. Here, an average treatment effect (ATE) reflects the sample average predicted change in internalizing/externalizing problems given a change in sleep hours. As causal decision trees can efficiently model high-order interaction terms, this method also uncovers the features contributing to heterogeneity in treatment effects. These conditional average treatment effects (CATE) reflect how treatment outcomes vary by observable characteristics of an individual. In doing so, we can parse heterogeneity to understand *for whom* a specific treatment would be more or less beneficial.

## METHODS

### Data

The ABCD study is a longitudinal project with 21 sites in the United States participating in data acquisition (abcdstudy.org). Starting at ages 9-10, biological, behavioral, and psychosocial information on children and their families are collected periodically as they advance into early adulthood (age 19-20). Parents provided written informed consent prior to participation, and children provided assent before data collection. Details on the ABCD study have been extensively described elsewhere^11,16^. Prior to running any analyses here, this study was partially pre-registered (from a larger study on prenatal cannabis exposures) on the open science framework platform (https://osf.io/a23s5).

Prenatal cannabis exposure was measured via a parent or caregiver self-report developmental history (‘DevHx’) questionnaire. Although individuals were asked about cannabis use before and after knowledge of their pregnancy, we collapsed across these exposure statuses to increase power to detect small effects. Likewise, a similar question was asked for any alcohol or tobacco use during their pregnancy. For all drug use questions, the parent or caregiver was asked to report about the biological mother’s use of drugs during their pregnancy.

Average sleep duration (in hours) was measured via the self-reported Sleep Disturbance Scale (SDS) for Children^17^. While sleep duration was collected on an ordinal scale (e.g., “5-7 hours”), the median at each interval was used for baseline and year-1 follow up. The difference between year-1 and baseline was calculated (so positive values signify an increase in sleep over time) and modelled as the treatment measure.

Child psychopathology was measured via the child behavior checklist (CBCL)^18^, which includes a summary T-score for total internalizing and externalizing problems, where higher scores reflect more problems. Likewise, the difference in problem scores between year-1 and baseline was calculated (i.e., positive values indicate an increase in problems over time) and used as the outcome measure. In light of the results from Paul et al.^12^ that found that children with prenatal cannabis exposure exhibited elevations in both the internalizing and externalizing domain, we ran two separate models (change in internalizing or externalizing problems as the outcome).

To account for confounding effects, key variables thought to be related to both sleep duration and internalizing/externalizing problems were included in the model. Specifically, sociodemographics like household income, parental education level, cohabitation status (i.e., married or separated), maternal age during pregnancy, child age, body-mass index, sex, and an indicator for white or non-white ethnicity were included. Other data from the mental health domain like maternal history of depression, and child baseline internalizing and externalizing scores were also included. Finally, any prenatal alcohol or tobacco use were also included as covariates.

Starting data were drawn from the baseline (N=11,873) and year-1 follow-up cohort contained in data release 3.0 from the ABCD study (abcdstudy.org). Participants were then filtered by having completed the developmental history questionnaire (n=11,533). While some siblings were included in ABCD, only one child from each family was randomly selected for inclusion to ensure independent sampling, thus yielding a sample of n=10,462. From there, only those with SDS and CBCL data available at baseline and year-1 were included for analysis (n=9,826). See figure 1 for inclusionary flow chart.

**Figure 1:**
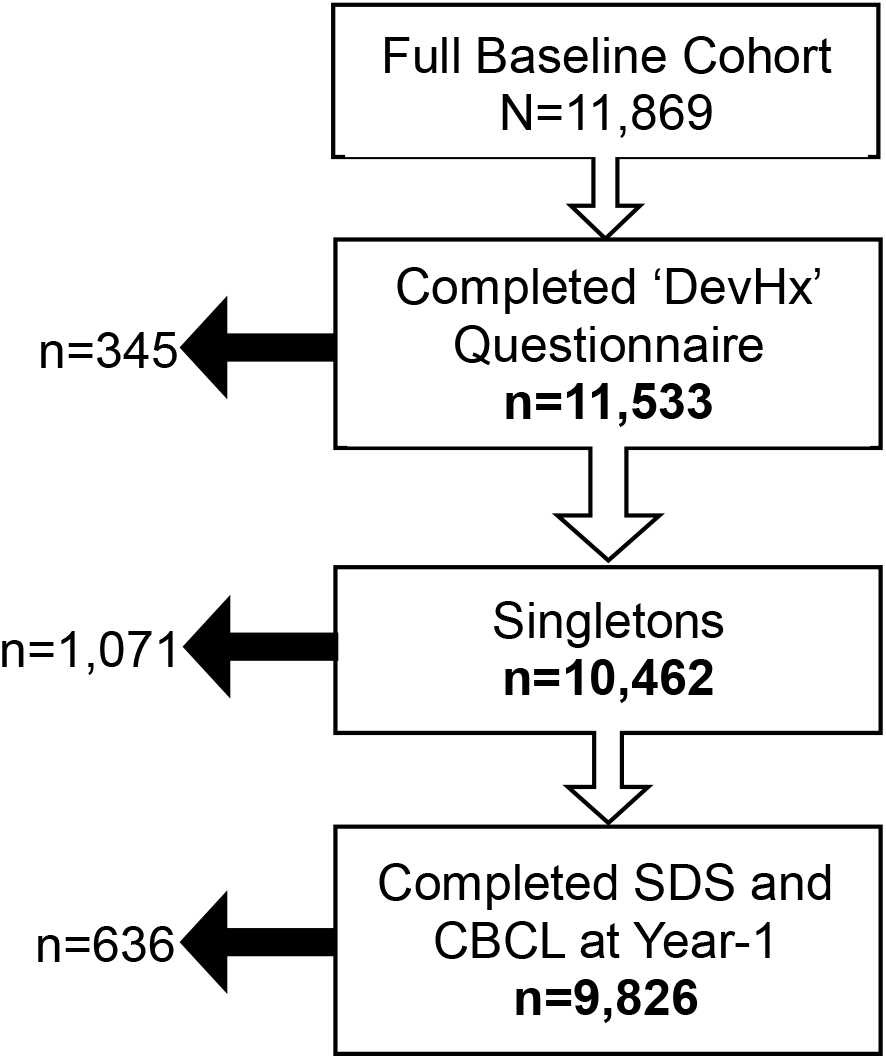
Participant inclusionary criteria ‘DevHx’ refers to the developmental history questionnaire which asks about prenatal drug exposures. Singletons refers to the random selection of one child per family in the ABCD study. ‘SDS’ refers to the sleep disturbance scale questionnaire (treatment measure), and ‘CBCL’ refers to the child behavior checklist (outcome measure).

### Generalized (Causal) Random Forest

Data were analyzed using the ‘grf’ package in R via the ‘causal forest’ function with default parameter settings (https://grf-labs.github.io/grf/). A causal random forest^19^ (CRF) is a data driven method designed to estimate the average and heterogeneous (e.g., moderated) treatment effects for a candidate causal pathway. Like a traditional random forest, a CRF constructs many decision trees to model a treatment effect on an outcome measure given a set of covariates. This approach can identify features that contribute to heterogeneity in the treatment effect (called conditional average treatment effects, CATE). Summary statistics for the CATEs are obtained using a doubly-robust estimation procedure. CRFs produce asymptotically normal standard errors and meets other assumptions necessary to perform statistical inference^15,19^. Moreover, the CRF uses “honest” tree splitting and out-of-bag model evaluation to enhance generalizability (for full details, see Athey et al., 2019^19^). Here, we modelled the effect of a change in sleep hours on a change in internalizing/externalizing, given a set of covariates (prenatal cannabis exposure, plus confounders), with acquisition site included as a clustering variable. See figure 2 for visualization of the causal diagram for this study.

**Figure 2:**
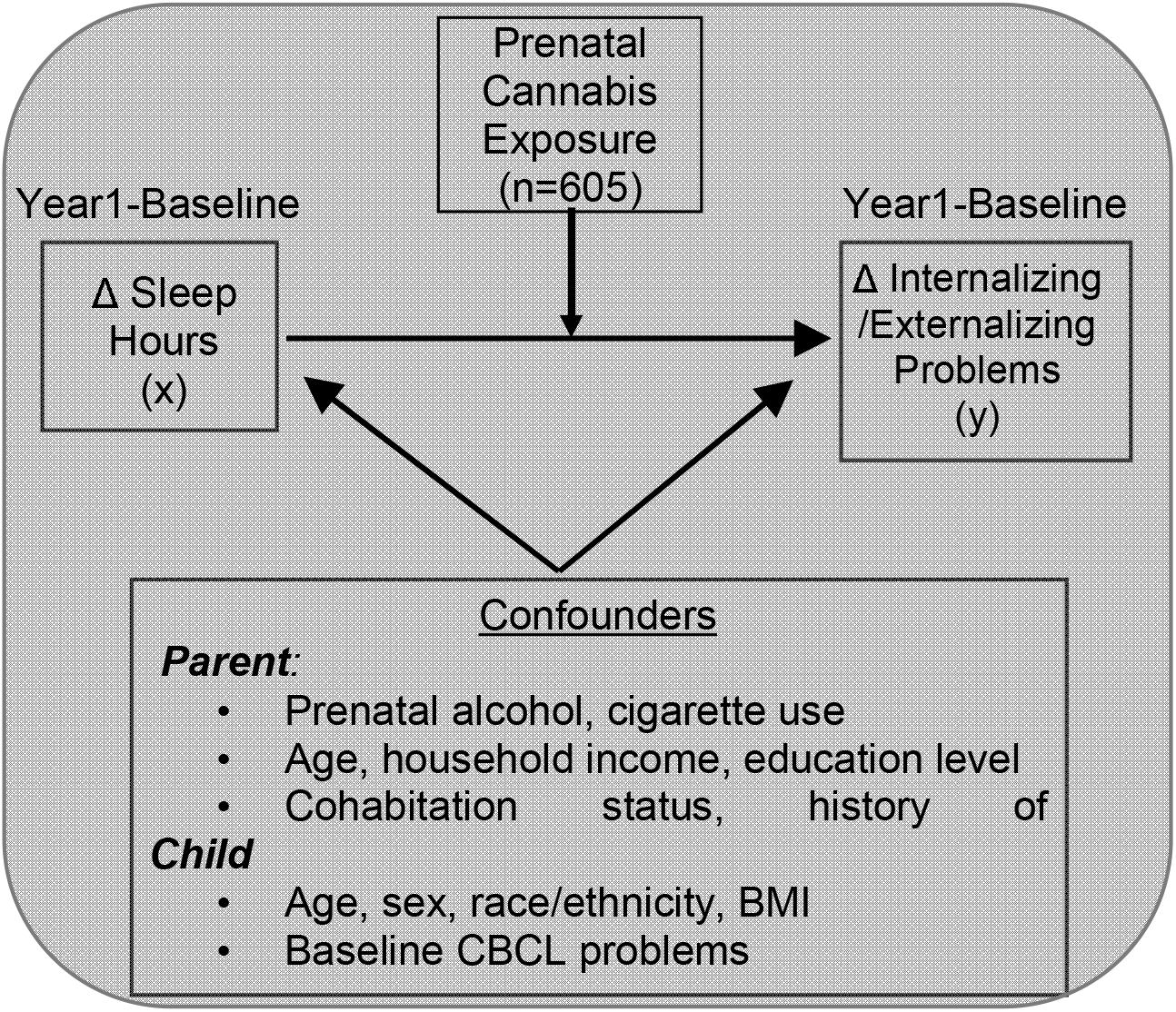
Causal diagram Conceptual model displaying that a change in sleep hours (treatment measure) was hypothesized to have a causal effect on a change in internalizing or externalizing problems (outcome measure). This causal pathway was hypothesized to be moderated by prenatal cannabis exposure status. A set of confounders related to both the treatment and outcome measures were included.

Any factors that were found to reflect significant conditional average treatment effects were probed further by plotting “simple slopes”^20^. For a binary factor (i.e., any prenatal cannabis exposure), the treatment effect of a change in sleep hours (x) on a change in internalizing/externalizing (y) was plotted at the two levels of the covariate (with vs. without exposure). Hence, this technique depicts the moderating effect of that covariate on the causal pathway and helps to convey the individual differences in treatment effects.

## RESULTS

From the full ABCD baseline cohort of 11,873 children, there were n=9,826 who met inclusionary criteria (figure 1) for analyses. Within that sample, there were n=605 with any prenatal cannabis exposure. See table 1 for information on the analytic sample, separated by prenatal cannabis exposure status for descriptive purposes.

Across the sample there was a slight decrease in sleep hours from baseline (*M*=9.01, *SD*=1.08) to year-1 (*M*=8.81, *SD*=1.13) (*t*_9,825_=-18.9, *p*<.001). Internalizing problems largely remained stable from baseline (*M*=48.54, *SD*=10.56) to year-1 (*M*=48.75, *SD*=10.64) (*t*_9,825_=2.40, *p*=.016). Externalizing problems also remained stable from baseline (*M*=45.64, *SD*=10.21) to year-1 (*M*=45.23, *SD*=10.09) (*t*_9,825_=5.44, *p*<.001).

**Table 1:** Sample demographics separated by prenatal cannabis exposure status. Table was divided by prenatal cannabis exposure (“CB Exposure”) status for descriptive purposes. P-values from two-sample t-tests (or Mann-Whitney *U* tests for categorical data). Note, given the large sample, small differences are significant at *p*<.05. All features except baseline sleep hours were included as covariates in causal random forest analyses.

| **Feature** | **No Exposure**<br>**(n=9,221)** | **CB Exposure**<br>**(n=605)** | ***p*** |
| --- | --- | --- | --- |
| Age in months (*M*, (*SD*)) | 118.80 (7.55) | 118.31 (7.64) | .12 |
| Sex (Female, %) | 4360 (47.3) | 303 (50.1) | .20 |
| Body Mass Index (mean (*SD*)) | 18.69 (3.95) | 19.82 (4.18) | <.01 |
| Race/Ethnicity (White, (%)) | 4969 (53.9) | 227 (37.5) | <.01 |
| Maternal Age of Pregnancy (mean ((*SD*)) | 29.70 (6.11) | 25.32 (6.07) | <.01 |
| Parents Married or Living Together (%) | 6978 (76.2) | 303 (50.6) | <.01 |
| Any Prenatal Alcohol Exposure (%) | 2115 (22.9) | 374 (61.8) | <.01 |
| Any Prenatal Tobacco Exposure (%) | 920 (10.0) | 362 (59.8) | <.01 |
| Maternal Depression (%) | 185 (2.0) | 137 (23.4) | <.01 |
| Household Income (%) |  |  | <.01 |
| <\$50k | 2345 (27.7) | 309 (55.8) | |
| >\$50k & <\$100k | 2421 (28.6) | 148 (26.7) | |
| >\$100k | 3693 (43.7) | 97 (17.5) | |
| Parental Highest Education (%) |  |  | <.01 |
| <HS Diploma | 461 (5.0) | 25 (4.1) |  |
| HS Diploma/GED | 799 (8.7) | 115 (19.0) |  |
| Some College | 2227 (24.2) | 266 (44.0) |  |
| Bachelors | 2386 (25.9) | 105 (17.4) |  |
| Post-Grad Degree | 3337 (36.2) | 94 (15.5) |  |
| BSL Internalizing Problems (mean (*SD*)) | 48.29 (10.47) | 52.28 (11.23) | <.01 |
| BSL Externalizing Problems (mean (*SD*)) | 45.25 (9.98) | 51.48 (11.80) | <.01 |
| BSL Sleep Hours (mean (*SD*)) | 9.03 (1.07) | 8.64 (1.21) | <.01 |

The GRF models uncovered an average treatment effect (ATE) for a change in sleep hours on a change in both internalizing (ATE=-.35, *SE*=.08, *p*=.003), and externalizing problems (ATE=-.28, *SE*=.07, *p*=.028). That is, for every 1 unit increase in sleep, the T-score decreased by .35 units for internalizing and by .28 units for externalizing, respectively.

For the internalizing model only, a conditional average treatment effect (CATE) was found by prenatal cannabis exposure (CATE=.92, *SE*=.36, *p*=.011). No other covariate contributed to a conditional average treatment effect for internalizing, and none were found for the externalizing model. See figure 3 (or supplementary table 1) for the CATE point estimates by each covariate, including prenatal alcohol and tobacco exposures, for the internalizing and externalizing models.

**Figure 3:**
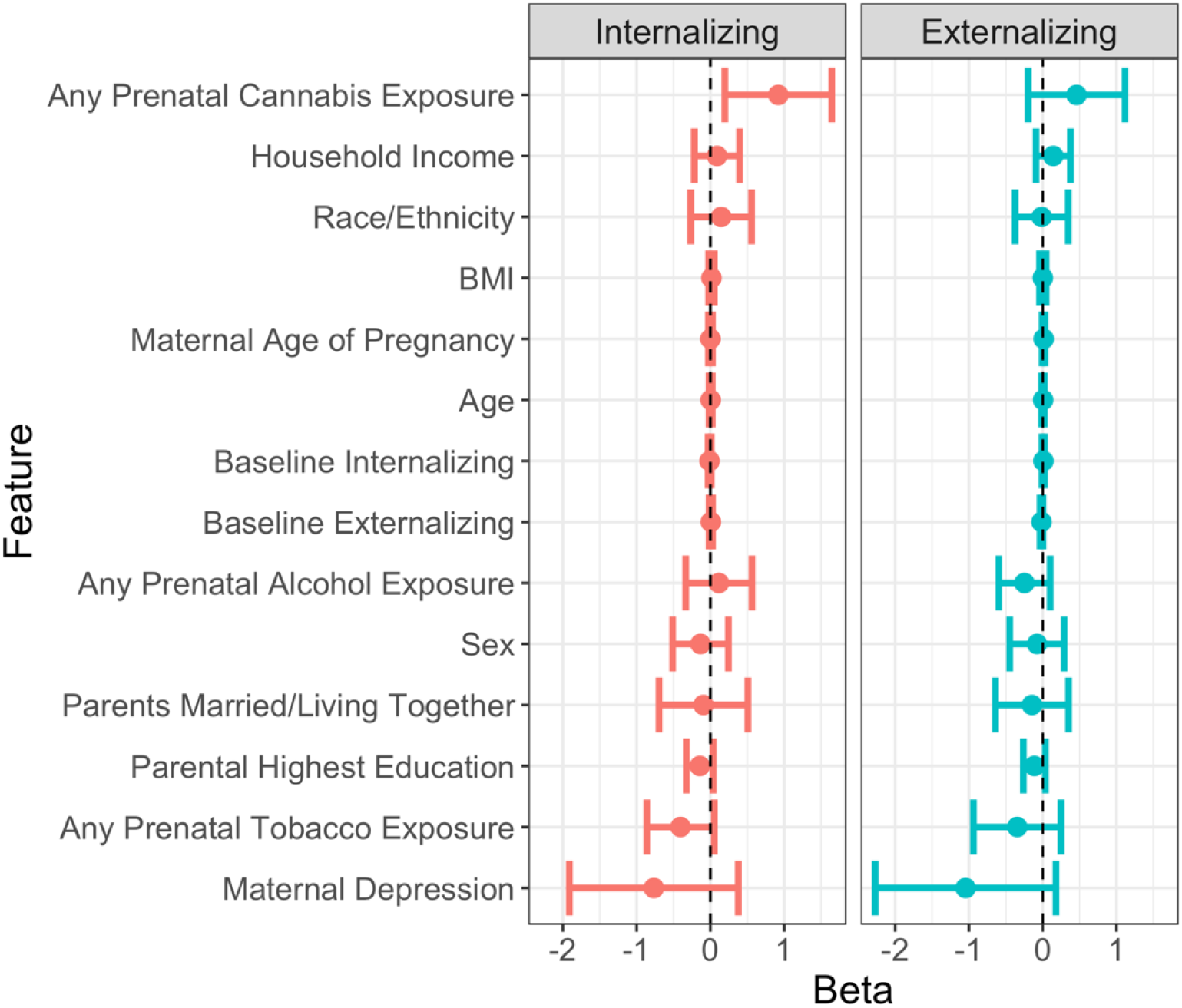
Conditional average treatment effects for the internalizing and externalizing models. Point estimates obtained from a doubly robust estimation procedure that regressed the outcome measure (internalizing/externalizing problems) onto the treatment measure (sleep hours) and covariates (tabulated here). Bars reflect two standard errors of the mean. The only significant conditional average treatment effect was found for changing sleep on changing internalizing problems by prenatal cannabis exposure.

To illuminate the nature of the identified conditional average treatment effect, simple slopes were plotted for children with vs. without prenatal cannabis exposure (figure 4). The average treatment effect was largely reproduced for children *without* prenatal cannabis exposure (*B*=-.38, *SE*=.08, n=9,221), while those *with* prenatal cannabis exposure exhibited divergent effects (*B*=.06, *SE=*.25, n=605), and the difference between those slopes was significant (*Z*=1.7, *p*_*one-sided*_ =.04). See supplementary figure 1 and 2 for similar plots separated by prenatal alcohol or tobacco exposure showing that participants with those exposures do not deviate from their non-exposed peers.

**Figure 4:**
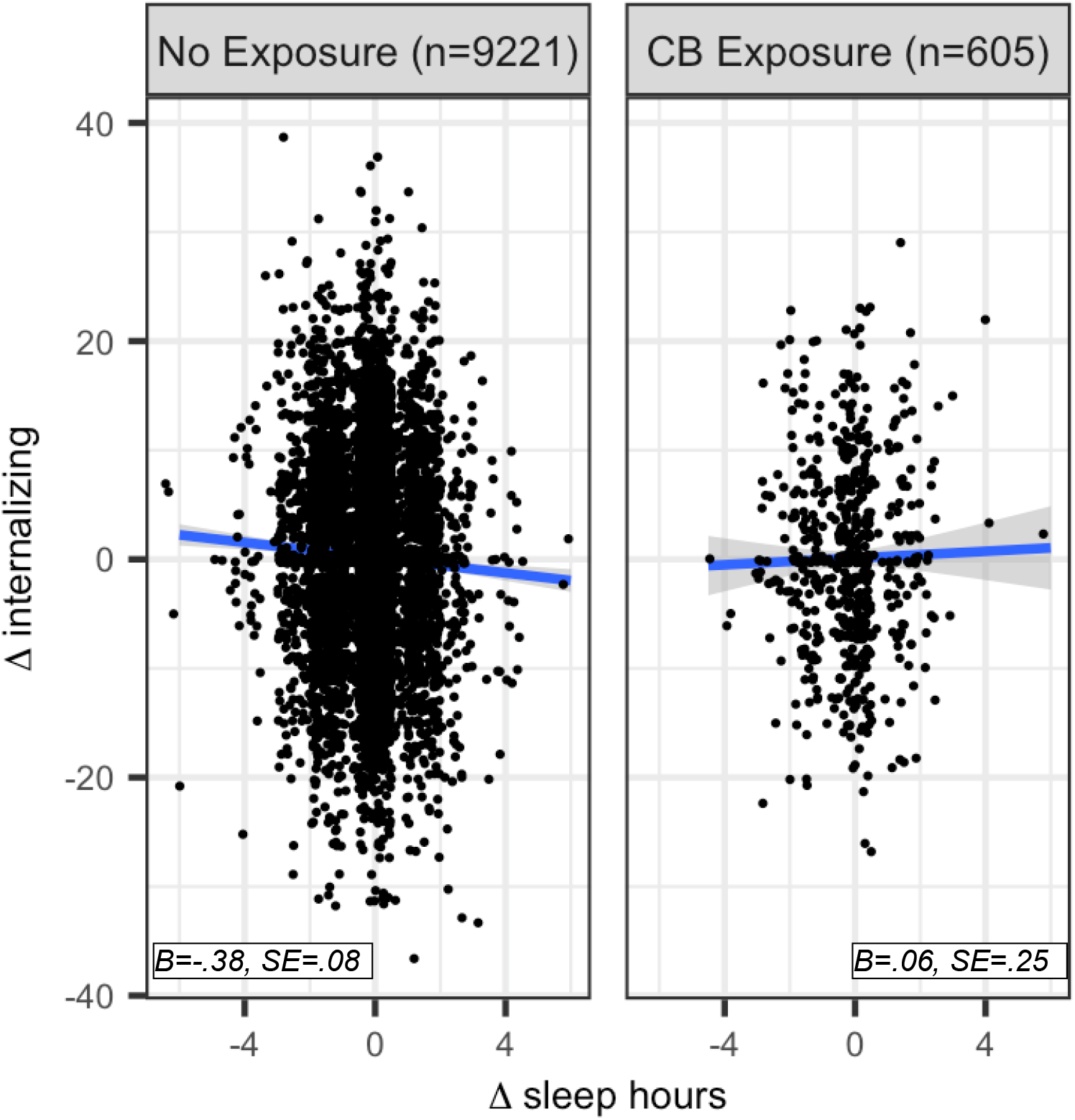
Conditional average treatment effect by prenatal cannabis exposure status on internalizing problems. Change in sleep hours (sleep disturbance scale) from baseline to year-1 follow up depicted on x-axis. Change in internalizing problems (child behavior checklist) from baseline to year-1 follow up depicted on y-axis. Separated by prenatal cannabis (“CB Exposure) status.

## DISCUSSION

This study examined whether the beneficial relationship between sleep and internalizing or externalizing problems was altered in children who were prenatally exposed to cannabis. Results indicated that an increase in sleep hours had a small causal effect on a decrease in internalizing and externalizing problems for the average child. This average treatment effect was moderated by prenatal cannabis exposure for internalizing problems, but not externalizing problems. Moreover, this finding appears to be specific to prenatal cannabis exposure, as heterogeneity was not found for prenatal alcohol or tobacco exposures. Hence, this study showcases how using a data driven causal inference framework can rigorously uncover substance use related individual differences in treatment effects.

The identified average treatment effect suggests that sleep is one modifiable behavioral domain that could be targeted to improve internalizing and externalizing problems for the average child. This finding is in line with other studies that found that sleep disturbances likely precede behavioral health problems in adolescence^3,4^. Therefore, building and maintaining healthy sleep behaviors could be a proactive way to protect against internalizing or externalizing problems in most typically developing children.

This study also suggests that certain subgroups of children (i.e., those with prenatal cannabis exposure) may not respond to sleep interventions in a similar fashion to their non-exposed peers. Hence, prenatal cannabis exposure might complicate treatment options as those children may require a different approach beyond sleep interventions to modify behavioral health problems. Nonetheless, these interpretations should be viewed considering the small effect sizes found here. And although small effects are to be expected^21,22^, they underscore how sleep and prenatal cannabis exposure are two elements that are part of a constellation of factors influencing mental health and wellbeing during child development.

These findings stand in contrast to the low risk perceptions^23^ and increasing prevalence of cannabis use during pregnancy. Data from a national survey indicated that 5.4% of pregnant individuals in 2019 reported any past-month cannabis use during their pregnancy. Recent evidence from a different sample found an increase in biochemically-verified cannabis use among pregnant individuals following the onset of the COVID-19 pandemic^24^ from 3.4 to 8.1% of individuals testing positive. As prenatal cannabis use becomes more prevalent, coupled with our findings that exposure may complicate mental health outcomes in children, research is needed to identify the specific facets of sleep, or other factors outside of the sleep domain, that can be modified in these children to achieve a similar benefit on their mood.

Future studies are also needed to determine the biological mechanisms that explain why and how prenatal cannabis exposure disrupts with the effect of sleep on mood. As the endocannabinoid system helps regulate neurodevelopment and sleep architecture^6^, there is a strong likelihood that prenatal cannabis exposures could interfere with those systems. This hypothesis is supported by the many studies reporting disadvantageous associations between prenatal cannabis use and child sleep features^7–10^. Hence, children with these exposures may be at a functional disadvantage to experience the same beneficial effects of sleep on their mood relative to their peers without such exposure. However, the precise mechanism linking endocannabinoid interference to mood-related outcomes remains unclear. Nonetheless, this study adds to the mounting evidence that prenatal cannabis use may impact the child’s brain and behavioral development and should thus motivate individuals to avoid using cannabis while pregnant.

Limitations of this study include the self-report nature of the treatment and outcome measures, which are not as reliable as objective measurement of sleep hours collected via wearable technology or biochemical verification of cannabis use during pregnancy. Additionally, the observational nature of the ABCD study, and the small effect sizes reported here should be considered when interpreting the possible causal nature of these findings. Despite using a rigorous causal inference framework, these results would benefit from being reproduced by a study using an experimental design that manipulates sleep hours. And although a set of confounders were included in the model, it is possible that there are omitted variables that could explain differences in sleep and behavior. Additionally, the use of two time points makes it difficult to parse out the temporal ordering of causal effects^25^. In future ABCD data releases, studies examining trajectories of sleep, mood, and behavior throughout adolescence will be possible by leveraging multi-timepoint data.

## Data Availability

All data produced in the present work are contained in the manuscript.

https://nda.nih.gov

## ACKNOWLEDGEMENTS

We thank Stefan Wager, Ph.D for helpful discussions regarding the analyses contained in this manuscript. The research conducted at the Laureate institute for Brain Research is supported by the William K. Warren Foundation. The ABCD Study is supported by the National Institutes of Health and additional federal partners under award numbers U01DA041022, U01DA041025, U01DA041028, U01DA041048, U01DA041089, U01DA041093, U01DA041106, U01DA041117, U01DA041120, U01DA041134, U01DA041148, U01DA041156, U01DA041174, U24DA041123, and U24DA041147.

