## Supplemental Table & Figures for "The beneficial effect of sleep on behavioral health problems in youth is disrupted by prenatal cannabis exposure: A causal random forest analysis of ABCD data"

**SUPPLEMENTARY MATERIAL**

Supplementary Table 1: Conditional average treatment effects for each model.

|  | **Internalizing Model** | | | **Externalizing Model** | | |
| --- | --- | --- | --- | --- | --- | --- |
| **Covariate** | ***B*** | ***Std.Err*** | ***p*** | ***B*** | ***Std.Err*** | ***p*** |
| Any Prenatal Cannabis Exposure | .92 | .36 | .01 | .46 | .33 | .16 |
| Any Prenatal Alcohol Exposure | .11 | .22 | .61 | -.25 | .17 | .15 |
| Any Prenatal Tobacco Exposure | -.40 | 23 | .08 | -.35 | .30 | .25 |
| Baseline Age | .003 | .01 | .76 | .006 | .01 | .55 |
| Sex | -.13 | .19 | .48 | -.08 | .19 | .68 |
| Body Mass Index | .01 | .02 | .44 | .002 | .02 | .94 |
| Race/Ethnicity | .15 | .21 | .48 | -.01 | .18 | .94 |
| Maternal Age of Pregnancy | .001 | .01 | .95 | .01 | .01 | .21 |
| Parents Married or Living Together | -.09 | .30 | .76 | -.15 | .25 | .56 |
| Maternal Depression | -.78 | .57 | .18 | -1.0 | .61 | .09 |
| Household Income | .09 | .15 | .56 | .14 | .12 | .22 |
| Parental Highest Education | -.14 | .09 | .12 | -.11 | .08 | .13 |
| BSL Internalizing Symptoms | -.01 | .01 | .36 | .01 | .01 | .32 |
| BSL Externalizing Symptoms | .006 | .01 | .61 | -.02 | .01 | .11 |

Point estimates obtained from a doubly robust estimation procedure that regressed the outcome measure (internalizing/externalizing problems) onto the treatment measure (sleep hours) and covariates (tabulated here). This data was visualized in figure 3.

Supplementary Figure 1: Conditional average treatment effect by prenatal alcohol exposure status on internalizing problems.

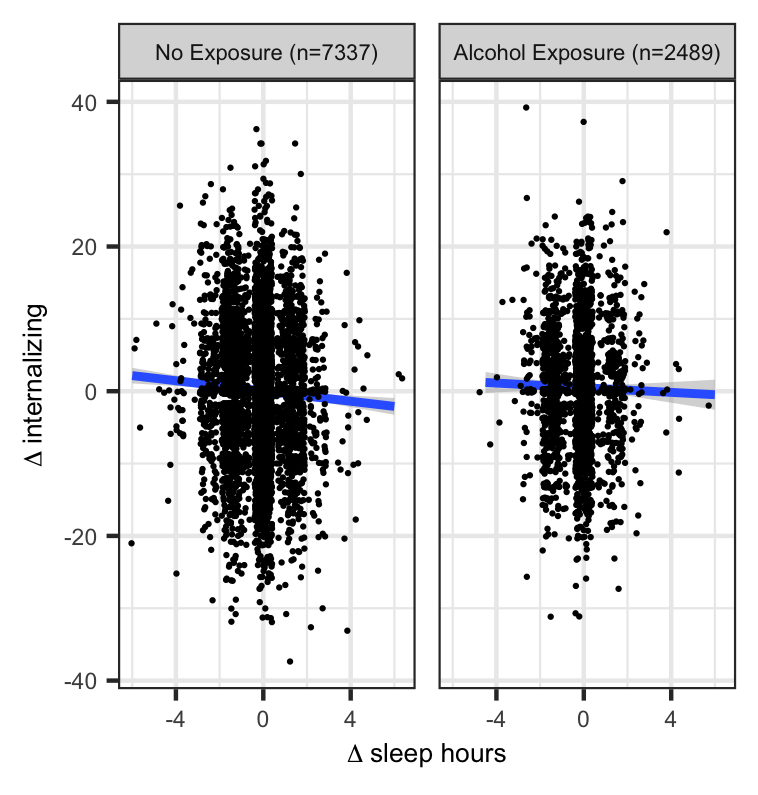

*B=-.39, SE=.10*

*B=-.24, SE=.16*

Change in sleep hours (sleep disturbance scale) from baseline to year-1 follow up depicted on x-axis. Change in internalizing problems (child behavior checklist) from baseline to year-1 follow up depicted on y-axis. Separated by prenatal alcohol exposure status.

Supplementary Figure 2: Conditional average treatment effect by prenatal tobacco exposure status on internalizing problems.

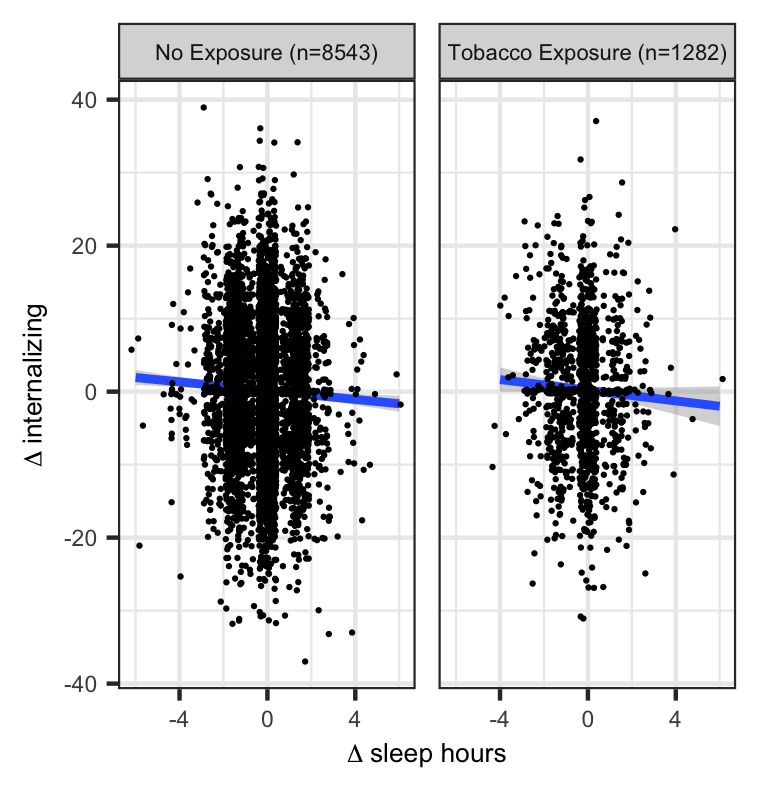

*B=-.33, SE=.09*

*B=-.56, SE=.15*

Change in sleep hours (sleep disturbance scale) from baseline to year-1 follow up depicted on x-axis. Change in internalizing problems (child behavior checklist) from baseline to year-1 follow up depicted on y-axis. Separated by prenatal tobacco exposure status.
